# An exploratory economic evaluation of the effect of the smokefree generation policy in England on smoking in pregnancy

**DOI:** 10.1101/2025.10.10.25337720

**Authors:** Nathan P Davies, Tessa Langley, Rachael L Murray, Joanne R Morling, Manpreet Bains, Matthew Jones

**Affiliations:** Nottingham Centre for Public Health and Epidemiology, Unit of Lifespan and Population Health, School of Medicine, University of Nottingham, UK; Centre of Academic Primary Care, Unit of Lifespan and Population Health, School of Medicine, University of Nottingham, UK; NIHR Nottingham Biomedical Research Centre, Nottingham University Hospitals NHS Trust with the University of Nottingham, UK

## Abstract

**Aims:** To estimate the impact of England’s proposed smokefree generation (SFG) policy on smoking at time of delivery (SATOD), adverse maternal and offspring outcomes, and the cost-effectiveness of the policy compared with no intervention.

**Design:** Decision analytic modelling using the Economics of Smoking in Pregnancy (ESIP) model, taking a health service and personal social services perspective. Three scenarios (central, pessimistic, optimistic) were simulated deterministically and through probabilistic sensitivity analyses (10,000 iterations).

**Setting:** England, United Kingdom.

**Participants:** A synthetic birth cohort of 19,843 women projected to give birth in 2042 at a mean maternal age of 28, representing the women affected by the first 15 years of the proposed 2027 SFG policy.

**Intervention and comparator:** The intervention is England’s proposed SFG policy to ban tobacco sales to those born on or after 1 January 2009. The comparator is no change in policy.

**Measurements:** The primary outcome is incremental cost-effectiveness ratio (ICER) per quality-adjusted life year (QALY) gained for combined maternal lifetime and offspring outcomes to age 15. Secondary outcomes are maternal and offspring outcomes (analysed separately), reductions in adverse pregnancy and offspring outcomes, life years gained, and benefit-cost ratio.

**Findings:** Across all scenarios, SFG dominated the comparator, with projections of both cost savings to health and personal social services and QALY gains. In the central scenario, SFG was projected to save 338 GBP and gain 0.152 QALYs per mother and child combined. The policy was estimated to avert 18 stillbirths, 58 premature births and 260 low-birthweight infants in 2042, and reduce child asthma prevalence by 0.8 percentage points and child mortality by 0.7 percentage points at age 15. When controlling for uncertainty, this dominance persisted in probabilistic sensitivity analyses, with all three scenarios having a 100% chance of being cost-effective. Additional analyses assuming 15-fold increased costs for each of the three scenarios still found SFG to be dominant.

**Conclusions:** In England, introducing a smokefree generation policy is projected to reduce smoking in pregnancy, prevent adverse birth outcomes, and deliver substantial health gains for mothers and children while reducing healthcare costs.

## INTRODUCTION

A smokefree generation (SFG) policy, also known as the tobacco-free generation policy, involves prohibiting tobacco sales to all individuals born after a certain point in time.[1] This is intended to slow uptake of tobacco use in new generations. The policy does not prohibit sales to those already dependent on nicotine, recognising the need for continued support to quit, but instead seeks to redefine tobacco as an unacceptably harmful product whose use should no longer be normalised.[2,3] Simulation studies for introducing SFG have been reported for New Zealand,[4,5] Singapore,[6,7] Solomon Islands,[8] Canada[9] and England[10]. They project the policy will lead substantial benefits in reducing smoking rates, and consequently tobacco-induced death and disease. However, most of these modelled health benefits take several decades to accrue, as people who were children or not even born at the time of policy implementation reach older age.

Women in pregnancy and their offspring may be exceptions for whom benefits could manifest much earlier. Smoking during pregnancy is a leading cause of adverse outcomes for both mother and child, including stillbirth[11], low birth weight[12], sudden infant death syndrome[11] and ectopic pregnancy.[13] Significant benefits could accrue in the immediate years and decades following introduction of a smokefree generation policy, when women affected by SFG begin giving birth.

Raising the age of sale of tobacco has empirically been linked to reductions in smoking in pregnancy. Tobacco 21 laws in the United States, which have likely reduced smoking prevalence in those aged under 21,[14] have also been shown to reduce smoking in pregnancy.[15,16] One study identified that this is likely driven by reduced smoking before pregnancy, rather than increased quitting during pregnancy;[15] another found evidence of a 9.5% reduction in incidence of adverse birth outcomes.[17]

Most SFG modelling studies have not included the impact on smoking in pregnancy, although it is an important short-term consideration for jurisdictions planning on introducing SFG. The United Kingdom (UK) government is seeking to introduce a SFG policy that would ban sales of tobacco (but not e-cigarettes) to those born after 1 January 2009.[18] If introduced, this would make the UK the second nation state, potentially following the Maldives in November 2025, to introduce such a policy.[19] The rate of smoking in pregnancy at time of delivery (SATOD) for England was 7.4% in 2023-2024[20] – comparatively low internationally. However, continued falls in prevalence are not guaranteed and there is a strong socioeconomic gradient, with the SATOD rate in the most deprived decile of postcodes almost twice as high as the England mean.[20] Studying the impact of SFG on pregnancy and childbirth could help decision-makers understand some of the earlier implications of introducing an SFG policy; indeed, the UK government identified a major limitation of their own modelling was that it did not take into account the effect on smoking in pregnancy.[18] We thus sought to explore the cost-effectiveness of SFG in England for reducing smoking rates during pregnancy.

## METHODS

### Decision analytic model

We used the Economics of Smoking in Pregnancy (ESIP) model to estimate incremental costs and health outcomes. ESIP is a peer reviewed decision analytic model that estimates the impact of changes in smoking both during and after pregnancy on both the health of the mother and her offspring. Further details of ESIP’s design and validation are reported elsewhere.[21,22]

We programmed ESIP with the following information: cohort size, year of pregnancy, age of mother, quit rates in comparator and intervention groups, and costs in comparator and intervention groups. All other ESIP inputs were unchanged. Analytical inputs are summarised in Table 1 and described in more detail below.

**Table 1:** Analytical inputs for ESIP modelling of SFG policy in England.

| Category | Parameter/scenario | Value | SE | Units | Source |
| --- | --- | --- | --- | --- | --- |
| **SATOD prevalence** | Comparator (no intervention) (2042) | 0.0652 | 0.00138 | Probability | [10] |
| **SATOD prevalence** | Pessimistic SFG (2042) | 0.0564 | 0.00113 | Probability | [10] |
| **SATOD prevalence** | Central SFG (2042) | 0.0493 | 0.00098 | Probability | [10] |
| **SATOD prevalence** | UK-gov SFG (2042) | 0.0293 | 0.00071 | Probability | [10] |
| **Demography** | Cohort size (live births) | 19,843 | — | Count | [10,23,24] |
| **Demography** | Mean maternal age at delivery | 28 | — | Years | [20] |
| **Cost** | Overall cost (comparator) | £0 | — | 2024 GBP | [18] |
| **Cost** | Overall cost (all SFG scenarios) | £1,500,000 | — | 2024 GBP | [18] |

### Cohort definition

We defined a single birth year cohort of females who would have been affected by the first 15 years of the UK’s SFG policy. The policy is due to come into force in 2027 and relates to those born in 2009 or later.[18] We assumed a mean maternal age at delivery of 28[20] leading to selection of females born in 2014. The total cohort was derived by multiplying the number of women aged 28 forecast to be in the UK in 2042[23] by the 6.45% of women forecast to smoke at time of delivery in 2042 (details below) by the 85% of women who have at least one child in their childbearing years.[24] This gave a cohort size of 19,843 women.

### Interventions

The intervention was introduction of a smokefree generation policy on 1 January 2027, affecting everyone born on 1 January 2009 or later.

We simulated one primary scenario and two secondary scenarios in which smokefree generation policies have differential effects on SATOD. These estimates of effect are based on prior synthesis of existing data on age-of-sale policies.[10] The scenarios modelled were:

(1) The central scenario, in which SFG results in a fixed drop in initiation rates and an additional small cumulative annual drop in initiation rates of 5%. This was estimated to reduce smoking at delivery to 4.93% (SE 0.098). (2) A pessimistic scenario analysis, in which SFG results in a fixed drop in initiation rates across all ages. This was estimated to reduce smoking at delivery to 5.64% (SE 0.113). (3) An optimistic scenario analysis, in which SFG results in a cumulative annual drop of initiation rates of 30%. This reduced smoking at delivery to 2.93% (SE 0.070).

ESIP was programmed with these cessation rates. Rather than treating SFG effects as increased quitting during pregnancy, we adjusted pre-pregnancy smoking status inputs so that the simulated share of women who smoke at delivery matched the scenario-specific SATOD levels. This approach treats SFG primarily as a reduction in smoking initiation before pregnancy while holding within-pregnancy cessation processes unchanged. Each SFG scenario was compared to the comparator scenario in which no additional women had quit in the usual policy environment.

### Comparator

The comparator was no change in policy, and therefore no change in the number of women smoking at delivery. We used estimates from an existing microsimulation model, designed to estimate the effects of the SFG policy on an English population, for prevalence of smoking amongst 28-year-old women giving birth in 2042. The model is described in full detail elsewhere.[10] Females in the least deprived Index of Multiple Deprivation quintile were used as a proxy for women smoking in pregnancy; this subgroup most closely approximates observed SATOD prevalence at the national level, thereby providing a reasonable benchmark for baseline smoking at delivery. The prediction was that if there was no SFG policy, 6.52% of women would be smoking at delivery. The comparator was also estimated to cost £0 per woman as there would be no additional policy cost.

### Estimating intervention costs

Estimated costs from a healthcare perspective are sourced from the Government’s impact assessment for the Tobacco and Vapes Bill, which estimates one-off costs for the Department of Health and Social Care (DHSC) to communicate the SFG policy. The population of women smoking in pregnancy to whom this cost will apply up to 2042 is assumed to be 15 times larger than the intervention population, because the communications cost is estimated to be one-off rather than annual.[18] The estimated one-off communication cost of introducing the policy was £1.5 million, giving a cost per woman of £5.04.

### Analytical strategy

We adopted an NHS and Personal Social Services perspective, with costs and benefits discounted at 3.5% per annum, as per NICE guidelines. [25] We focused on costs and outcomes for the mother up to the age of 100 years. It is likely that the SFG policy would have large impacts on the offspring’s smoking behaviour[10], and therefore we would anticipate large changes in the probabilities for offspring initiating smoking that are separate from the effect of changes in maternal smoking. Given these are likely to be very different from the original ESIP inputs, we took a conservative approach for the offspring and estimated impacts up to the age of 15 years rather than across the lifespan.

Our primary measure of cost-effectiveness was the Incremental Cost-Effectiveness Ratio (ICER) per Quality-Adjusted Life Year (QALY) gained for combined maternal and offspring outcomes. Our secondary measures of cost-effectiveness were ICERs per QALY gained for maternal and infants separately, ICERs per Life Year (LY) gained, and Return on Investment (ROI) for reductions in healthcare costs only. We also estimated the impact on adverse maternal and infant birth outcomes, and offspring mortality and morbidity at age 15 years.

### Characterising uncertainty

To quantify the impact of uncertainty, we carried out a probabilistic sensitivity analysis (PSA), allowing all ESIP inputs to vary within pre-defined ranges. Given the large number of parameters, 10,000 Monte Carlo simulations[26] were run to estimate incremental costs and benefits for each pairwise comparison. PSA results were plotted through cost-effectiveness acceptability curves (CEACs) and scatterplots.[27] As an additional scenario analysis, we also explored an alternative costing assumption in which the one-off government communication cost was fully allocated to each single-year pregnancy cohort (i.e. 15 times higher per woman than in the three main SFG scenarios).

### Approach and effect of engaging with PPI

For the overarching research programme on the smokefree generation, three groups of 12-to 21-year-olds in England (supported in line with NIHR guidance[28]) provided views on important outcome measures for studying the smokefree generation policy, which included cost savings to the NHS and future health outcomes, leading to focus on economic modelling.

## RESULTS

### Health economic outcomes

All three scenarios suggested that the SFG intervention was dominant; in all cases, the SFG intervention was found to simultaneously reduce healthcare costs and lead to increases in QALYs and LYs (Table 2). The pessimistic scenario produced the smallest gains in QALYs and LYs, but still recorded a ROI of £35.14 for combined maternal and offspring outcomes, meaning that for £1 spent on SFG, a return of £35.14 in healthcare savings was estimated. The biggest gains in QALYs and healthcare savings in all scenarios were amongst the offspring. However, SFG was still dominant in all scenarios when focusing on maternal outcomes only.

**Table 2:** Base case findings for primary incremental outcomes and incremental cost-effectiveness ratios (ICERs) for maternal lifetime, infant childhood up to age 15, and combined mother and infant, across three scenarios.

|  | Central SFG scenario |  | Pessimistic SFG scenario |  | Optimistic SFG scenario |  |
| --- | --- | --- | --- | --- | --- | --- |
| Outcome and time horizon | Incremental | ICER (£) | Incremental | ICER (£) | Incremental | ICER (£) |
| **Expected cost (£)** |  |  |  |  |  |  |
| Mother | -126.40 |  | -65.50 |  | -298.96 |  |
| Offspring | -206.21 |  | -108.32 |  | -483.54 |  |
| Combined | -337.64 |  | -178.86 |  | -782.55 |  |
| **LYGs** |  |  |  |  |  |  |
| Mother | 0.017 | -7290.16 | 0.0093 | -7039.18 | 0.040 | -7455.17 |
| Offspring | 0.086 | -2404.90 | 0.046 | -2354.15 | 0.198 | -2438.27 |
| Combined | 0.103 | -3275.49 | 0.055 | -3233.27 | 0.238 | -3303.24 |
| **QALYs** |  |  |  |  |  |  |
| Mother | 0.054 | -2345.31 | 0.029 | -2264.57 | 0.125 | -2398.40 |
| Offspring | 0.098 | -2099.41 | 0.052 | -2055.10 | 0.227 | -2128.53 |
| Combined | 0.152 | -2219.66 | 0.081 | -2191.05 | 0.352 | -2238.47 |
| **Benefit-cost ratio** |  |  |  |  |  |  |
| Mother |  | 26.08 |  | 14.00 |  | 60.32 |
| Offspring |  | 41.91 |  | 22.49 |  | 96.94 |
| Combined |  | 65.48 |  | 35.14 |  | 151.44 |

### Health outcomes

In the central scenario, SFG averted an average of 109 adverse pregnancy events and 121 adverse live births per 19,843 pregnancies for women giving birth in 2042. Reductions were also observed in stillbirths (18 fewer), premature births (58 fewer), and low-birthweight infants (260 fewer). By age 15, the prevalence of offspring asthma fell by 0.8 percentage points, and child mortality by 0.7 percentage points. Again, the pessimistic and optimistic scenarios produced smaller and larger benefits respectively, but direction of effect was consistent. Results are presented in Table 3.

**Table 3:** Base case findings for secondary incremental outcomes for maternal lifetime, infant childhood up to age 15, and combined mother and infant, across three scenarios.

|  | **Central SFG scenario** | **Pessimistic SFG scenarios** | **Optimistic SFG scenario** |
| --- | --- | --- | --- |
| **Outcome and time horizon** | **Incremental** | **Incremental** | **Incremental** |
| **Number of adverse pregnancy events** | -109 | -58 | -251 |
| **Number of stillbirths** | -18 | -10 | -43 |
| **Number of infants born prematurely** | -58 | -31 | -133 |
| **Number of infants born with LBW** | -260 | -140 | -601 |
| **Number of adverse live births** | -121 | -65 | -279 |
| **Number of any adverse infant outcomes** | -268 | -144 | -620 |
| **Proportion of infants dead (age 15)** | -0.74% | -0.40% | -1.72% |
| **Proportion of infants with asthma (age 15)** | -0.80% | -0.42% | -1.82% |

### Probabilistic sensitivity analysis (PSA)

Results from the PSA, based on 10,000 Monte Carlo simulations, confirmed the robustness of base case findings. In all simulations, each SFG scenario dominated the no-intervention comparator, meaning that it was always estimated to be cost-saving and lead to an increase in QALYs and LYs. For the central scenario, combined maternal and offspring cost savings were estimated at £344 (95% CI £242 – £441) per pregnancy, with incremental QALY gains of 0.154 (95% CI: 0.104 - 0.234). The benefit-cost ratio for the combined population was 67 (95% CI: 43 - 95). Full PSA estimates are shown in Table 4.

**Table 4:** Probabilistic primary and secondary incremental outcomes and incremental cost-effectiveness ratios (ICERs) for maternal lifetime, infant childhood up to age 15, and combined mother and infant, for three SFG scenarios vs comparator.

| Central SFG scenario |  |  |
| --- | --- | --- |
| Outcome and population | Incremental | ICER (£)/BCR |
| **Expected cost (£)** |  |  |
| Mother | -130.08 (-183.95, -83.74) |  |
| Offspring | -209.15 (-297.81, -120.42) |  |
| Combined | -344.28 (-449.53, -241.64) |  |
| **LYGs** |  |  |
| Mother | 0.02 (0.01, 0.03) | -7,598.59 (-11,856.28, -4,745.82) |
| Offspring | 0.09 (0.06, 0.12) | -2,497.70 (-4,103.40, -1,259.71) |
| Combined | 0.10 (0.08, 0.13) | -3,389.69 (-5,091.64, -2,084.80) |
| **QALYs** |  |  |
| Mother | 0.05 (0.03, 0.09) | -2,589.67 (-4,939.34, -1,389.52) |
| Offspring | 0.10 (0.06, 0.17) | -2,251.17 (-3,916.94, -990.70) |
| Combined | 0.15 (0.10, 0.23) | -2,329.89 (-3,596.19, -1,338.65) |
| **Benefit-cost ratio** |  |  |
| Mother |  | 27.04 (17.03, 39.90) |
| Offspring |  | 42.38 (23.29, 63.35) |
| Combined |  | 66.90 (42.66, 94.99) |
| **Pessimistic SFG scenario** |  |  |
| **Expected cost (£)** |  |  |
| Mother | -66.94 (-95.60, -42.33) |  |
| Offspring | -109.92 (-159.58, -61.22) |  |
| Combined | -181.91 (-239.85, -125.97) |  |
| **LYGs** |  |  |
| Mother | 0.01 (0.01, 0.01) | -7,312.08 (-11,310.12, -4,527.78) |
| Offspring | 0.05 (0.03, 0.06) | -2,437.65 (-4,043.22, -1,195.45) |
| Combined | 0.06 (0.04, 0.07) | -3,329.44 (-5,018.69, -2,023.10) |
| **QALYs** |  |  |
| Mother | 0.03 (0.01, 0.05) | -2,502.54 (-4,782.86, -1,328.39) |
| Offspring | 0.05 (0.03, 0.09) | -2,192.06 (-3,824.47, -956.36) |
| Combined | 0.08 (0.06, 0.13) | -2,291.35 (-3,575.01, -1,293.81) |
| **Benefit-cost ratio** |  |  |
| Mother |  | 14.40 (8.99, 21.30) |
| Offspring |  | 22.70 (12.43, 34.49) |
| Combined |  | 35.73 (22.53, 51.04) |
| **Optimistic SFG scenario** |  |  |
| **Expected cost (£)** |  |  |
| Mother | -306.15 (-433.72, -200.14) |  |
| Offspring | -490.04 (-696.77, -283.02) |  |
| Combined | -801.24 (-1,050.72, -562.17) |  |
| **LYGs** |  |  |
| Mother | 0.04 (0.03, 0.06) | -2,645.94 (-5,007.50, -1,423.10) |
| Offspring | 0.20 (0.14, 0.27) | -2,529.67 (-4,194.80, -1,257.61) |
| Combined | 0.24 (0.18, 0.31) | -3,409.58 (-5,160.66, -2,095.16) |
| **QALYs** |  |  |
| Mother | 0.13 (0.06, 0.21) | -7,730.96 (-11,765.73, -4,874.09) |
| Offspring | 0.23 (0.15, 0.40) | -2,282.58 (-3,960.23, -1,023.35) |
| Combined | 0.35 (0.24, 0.54) | -2,348.41 (-3,640.73, -1,364.86) |
| **Benefit-cost ratio** |  |  |
| Mother |  | 62.36 (38.96, 92.42) |
| Offspring |  | 97.95 (53.16, 146.67) |
| Combined |  | 154.43 (97.84, 218.83) |

### Alternative costing assumption

Results were robust to the alternative costing assumption (Supplementary Material T1). Even when intervention costs were increased 15-fold, the central scenario still showed dominance, with combined savings of £267 per pregnancy and incremental QALY gains of 0.15. The benefit-cost ratio fell from 65:1 in the base case to 4:1, but the policy remained highly cost-effective.

### Cost-effectiveness distributions

The cost-effectiveness acceptability curves for the central scenario, pessimistic scenario and optimistic scenario all showed a certainty of 1 for cost-effectiveness at all willingness-to-pay QALY thresholds up to a value of £100,000 (Figure 1). The scatterplot of incremental costs against incremental QALYs illustrates that all 10,000 iterations of the central scenario fell in the southeast quadrant of the cost-effectiveness plane, indicating both greater health benefit and lower costs compared to no intervention (Supplementary Material F1).

**Figure 1:**
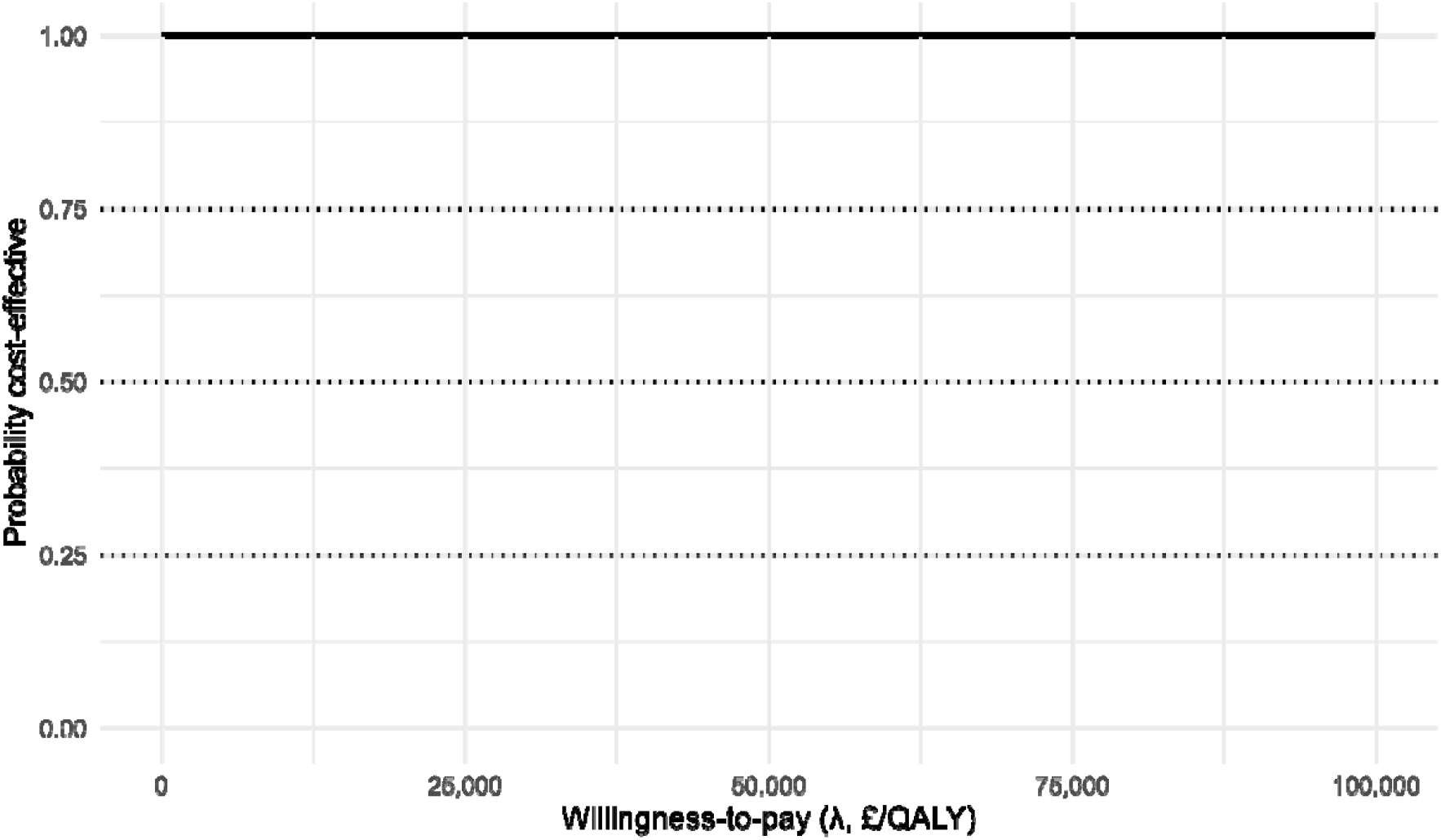
Cost-effectiveness acceptability curve for each of the three SFG scenarios compared to no SFG comparator.

## DISCUSSION

Our analyses indicate that the projected reduction in smoking at time of delivery leads to substantial health gains for mothers and infants and long-term savings for the NHS and social care. Across all scenarios, SFG dominated the no-intervention comparator, producing both cost savings and quality-of-life gains over the mother’s lifetime and the child’s early years. The central scenario indicated moderate per-pregnancy savings and QALY gains, with prevention of a notable number of adverse pregnancy outcomes. Benefits extended into childhood, with lower risks of asthma and mortality by age 15. Even under pessimistic assumptions and conservative costing, SFG consistently remained cost-effective, showing dominance across all willingness-to-pay thresholds and robustness to variation in government communication costs.

Our results align with empirical evidence from the United States, where raising the age of sale of tobacco to 21 (Tobacco 21) has been associated with reductions in smoking during pregnancy[15,16] and improvements in birth outcomes.[17] This strengthens confidence that reductions in smoking at delivery projected under SFG are plausible. Pregnancy is a life stage where benefits accrue more quickly: women in the first cohorts affected by SFG will reach childbearing age well before chronic smoking-related diseases typically manifest. Intergenerational pathways are also relevant; lower maternal smoking prevalence reduces the likelihood of smoking initiation among offspring[29] although these longer-term benefits were not modelled here.

Our findings highlight near-term benefits in pregnancy and early childhood, addressing a limitation identified in the UK government’s own economic modelling, which did not include smoking in pregnancy.[30] If even modest reductions in smoking initiation are achieved, SFG is likely to be a highly cost-effective intervention for improving maternal and child health, even before accounting for the larger downstream gains in adult disease reduction. The health gains reported here are a snapshot for a single year cohort; in reality, should smoking initiation reduce as projected, health benefits will continue to accumulate over decades. These findings suggest that governments should view SFG not only as a population-level prevention policy but also as a maternal and child health intervention. Even greater gains may be possible for nations with higher smoking rates in pregnancy; for example, in France, mothers of 0–5-year-olds reported the same levels of smoking in their pregnancy (13%) between 2017 and 2021.[31] However, nations with higher youth smoking rates and poor adherence to existing age-of-sale policies may be more likely to find it difficult to secure political and societal agreement for, and to enforce, an SFG policy.[2]

If implemented in England, monitoring trends in smoking initiation among young people will provide the earliest indication of policy impact. Should SFG successfully reduce initiation, it is reasonable to expect subsequent reductions in smoking at time of delivery, with associated health and cost benefits emerging over time. Further modelling could explore health and cost outcomes for smoking in pregnancy for other settings considering implementation of an SFG policy.

### Strengths and limitations

This is the first study to quantify the impact of SFG on smoking in pregnancy and related outcomes, using the validated Economics of Smoking in Pregnancy (ESIP) model. By linking a population-level tobacco model to a pregnancy-specific model, we capture outcomes that other SFG models have not considered. The analysis also considered maternal lifetime horizons alongside offspring outcomes to age 15, demonstrating sizeable benefits at multiple life stages.

There are some limitations to our study. First, we used smoking rates among women in the least deprived quintile as a proxy for smoking at delivery, which may not fully capture socioeconomic variation in smoking during pregnancy. Second, we modelled only a single cohort year, meaning that aggregate benefits across successive years of the policy will be considerably larger. Third, we adopted a conservative approach by excluding potential long-term impacts on offspring smoking initiation and adult disease risk. Fourth, we have used official government estimates of communication costs, but because of this study’s health and care perspective, these analyses do not consider any potential additional enforcement costs. However, we have assumed the costs of the policy only apply to pregnant women who would have initiated smoking, when in fact they will apply to the entire population, thus overestimating their impact. Finally, possible substitution to e-cigarettes was not modelled. Taken together, this study likely underestimates the true health and economic benefits of SFG from a health and care perspective.

## CONCLUSIONS

Our analyses suggest that SFG is likely to deliver substantial early benefits for pregnancy and childhood outcomes in England, alongside long-term gains, even under pessimistic assumptions of effectiveness of SFG. For nations for whom SFG is a viable tobacco control policy, our strengthens the case for adoption of SFG as a cost-saving tobacco endgame policy.

## ACKNOWLEDGEMENTS

This study is funded by the HEE/NIHR Integrated Clinical Academic Programme (grant NIHR302872). The views expressed are those of the authors and not necessarily those of the NIHR or the Department of Health and Social Care.

## DECLARATIONS OF INTEREST

None.

## DATA AVAILABILITY STATEMETN

This article makes use of several openly available government and statistical agency sources, all of which are referenced with links. Details on the ESIP model are available at https://www.nottingham.ac.uk/research/groups/tobaccoandalcohol/smoking-in-pregnancy/esip/index.aspx. Full data is available on reasonable request.

## Notes

### Competing Interest Statement

The authors have declared no competing interest.

### Author Declarations

This article makes use of several openly available government and statistical agency sources of human data, all of which are referenced with links. Details on the ESIP model are available at https://www.nottingham.ac.uk/research/groups/tobaccoandalcohol/smoking-in-pregnancy/esip/index.aspx.

## References

[1] Berrick AJ. The tobacco-free generation proposal. Tob Control 2013;22:i22. 10.1136/tobaccocontrol-2012-050865.

[2] Berrick J. Guidance for Introducing the Tobacco-Free Generation Policy. Int J Health Plann Manage 2025. 10.1002/HPM.3896.

[3] Khoo D, Chiam Y, Ng P, Berrick AJ, Koong HN. Phasing-out tobacco: proposal to deny access to tobacco for those born from 2000. Tob Control 2010;19:355. 10.1136/tc.2009.031153.

[4] Ouakrim DA, Wilson T, Waa A, Maddox R, Andrabi H, Mishra SR, et al. Tobacco endgame intervention impacts on health gains and Māori:non-Māori health inequity: a simulation study of the Aotearoa/New Zealand Tobacco Action Plan. Tob Control 2023:tc-2022-057655. 10.1136/tc-2022-057655.

[5] Deen FS van der, Wilson N, Cleghorn CL, Kvizhinadze G, Cobiac LJ, Nghiem N, et al. Impact of five tobacco endgame strategies on future smoking prevalence, population health and health system costs: two modelling studies to inform the tobacco endgame. Tob Control 2018;27:278. 10.1136/tobaccocontrol-2016-053585.

[6] Zeng Z, Cook AR, Eijk Y van der. What measures are needed to achieve a tobacco endgame target? A Singapore-based simulation study. Tob Control 2023:tc-2022-057856. 10.1136/tc-2022-057856.

[7] Doan TTT, Tan KW, Dickens BSL, Lean YA, Yang Q, Cook AR. Evaluating smoking control policies in the e-cigarette era: a modelling study. Tob Control 2020;29:522. 10.1136/tobaccocontrol-2019-054951.

[8] Singh A, Petrović-Van Der Deen FS, Carvalho N, Lopez AD, Blakely T. Impact of tax and tobacco-free generation on health-adjusted life years in the Solomon Islands: a multistate life table simulation. Tob Control 2020;29:388–97. 10.1136/TOBACCOCONTROL-2018-054861.

[9] Coyle D. Implementing a smoke-free generation policy for Canada: estimates of the long-term impacts. Health Promot Chronic Dis Prev Can 2025;45:39–53. 10.24095/HPCDP.45.1.03.

[10] Davies NP, Murray RL, Morling JR, Bains M, Jones M, Langley T. Impact of the United Kingdom’s smokefree generation policy on tobacco-related equity in England: a simulation study. MedRxiv 2025:2025.06.13.25328563. 10.1101/2025.06.13.25328563.

[11] Pineles BL, Hsu S, Park E, Samet JM. Systematic Review and Meta-Analyses of Perinatal Death and Maternal Exposure to Tobacco Smoke During Pregnancy. Am J Epidemiol 2016;184:87–97. 10.1093/AJE/KWV301.

[12] Di HK, Gan Y, Lu K, Wang C, Zhu Y, Meng X, et al. Maternal smoking status during pregnancy and low birth weight in offspring: systematic review and meta-analysis of 55 cohort studies published from 1986 to 2020. World Journal of Pediatrics 2022;18:176–85. 10.1007/S12519-021-00501-5/TABLES/1.

[13] Avşar TS, McLeod H, Jackson L. Health outcomes of smoking during pregnancy and the postpartum period: an umbrella review. BMC Pregnancy Childbirth 2021;21:1–9. 10.1186/S12884-021-03729-1/TABLES/2.

[14] Davies N, Bogdanovica I, McGill S, Murray RL. What is the Relationship Between Raising the Minimum Legal Sales Age of Tobacco Above 20 and Cigarette Smoking? A Systematic Review. Nicotine & Tobacco Research 2024. 10.1093/NTR/NTAE206.

[15] Bersak T, Lavender M, Sonchak-Ardan L. Impact of Tobacco-21 Laws on Maternal Smoking Behavior. Health Econ 2025. 10.1002/HEC.4951.

[16] Tennekoon VSBW. Effects of Purchase Restrictions on Smoking During Pregnancy: An Analysis of U.S. Birth Records. Nicotine & Tobacco Research 2023;25:882–8. 10.1093/NTR/NTAC220.

[17] Tennekoon VSBW. Purchase restrictions as a tobacco control policy: An analysis of the effect on adverse birth outcomes. Econ Anal Policy 2023;78:967–74. 10.1016/J.EAP.2023.04.030.

[18] Department of Health and Social Care. The Tobacco and Vapes Bill: impact assessment 2024. https://www.gov.uk/government/publications/the-tobacco-and-vapes-bill-impact-assessment (accessed March 20, 2025).

[19] Tobacco Tactics. The Maldives passes landmark legislation introducing a generational ban on tobacco use 2025. https://www.tobaccotactics.org/news/the-maldives-passes-landmark-legislation-introducing-a-generational-ban-on-tobacco-use/ (accessed August 15, 2025).

[20] NHS England. Statistics on Women’s Smoking Status at Time of Delivery: Additional analysis, England, 2023-24 2024.

[21] University of Nottingham. Economics of Smoking in Pregnancy (ESIP) Model n.d. https://www.nottingham.ac.uk/research/groups/tobaccoandalcohol/smoking-in-pregnancy/esip/index.aspx (accessed September 4, 2025).

[22] Jones M, Smith M, Lewis S, Parrott S, Coleman T. A dynamic, modifiable model for estimating cost-effectiveness of smoking cessation interventions in pregnancy: application to an RCT of self-help delivered by text message. Addiction 2019;114:353–65. 10.1111/add.14476.

[23] Office for National Statistics. Principal projection - England summary 2025. https://www.ons.gov.uk/peoplepopulationandcommunity/populationandmigration/populationprojections/datasets/tablea14principalprojectionenglandsummary (accessed July 24, 2025).

[24] Office for National Statistics. How is the fertility rate changing in England and Wales? 2024. https://www.ons.gov.uk/peoplepopulationandcommunity/birthsdeathsandmarriages/conceptionandfertilityrates/articles/howisthefertilityratechanginginenglandandwales/2024-10-28 (accessed March 20, 2025).

[25] National Institute for Health and Care Excellence. NICE health technology evaluations: the manual 2023. https://www.nice.org.uk/process/pmg36/chapter/economic-evaluation-2#discounting (accessed March 20, 2025).

[26] Doubilet P, Begg CB, Weinstein MC, Braun P, Mcneil BJ. Probabilistic Sensitivity Analysis Using Monte Carlo Simulation: A Practical Approach. Medical Decision Making 1985;5:157–77. 10.1177/0272989X8500500205

[27] Fenwick E, Marshall DA, Levy AR, Nichol G. Using and interpreting cost-effectiveness acceptability curves: An example using data from a trial of management strategies for atrial fibrillation. BMC Health Serv Res 2006;6:1–8. 10.1186/1472-6963-6-52.

[28] National Institute for Health and Care Research. Public involvement in research. 2024 n.d. https://www.nihr.ac.uk/get-involved/public-involvement (accessed October 9, 2025).

[29] Leonardi-Bee J, Jere ML, Britton J. Exposure to parental and sibling smoking and the risk of smoking uptake in childhood and adolescence: a systematic review and meta-analysis. Thorax 2011;66:847–55. 10.1136/THX.2010.153379.

[30] Department of Health and Social Care. Modelling for the smokefree generation policy 2023. https://www.gov.uk/government/publications/smokefree-generation-policy-modelling-report/modelling-for-the-smokefree-generation-policy (accessed March 8, 2024).

[31] Santé publique France. Tobacco and alcohol use during pregnancy. Results of the 2021 French Public Health Barometer. 2024.

