## Supplementary material for "An exploratory economic evaluation of the effect of the smokefree generation policy in England on smoking in pregnancy"

#### Supplementary Table 1: Sensitivity analysis findings for 15-fold increased costs for primary incremental outcomes and incremental cost-effectiveness ratios (ICERs) for maternal lifetime, infant childhood up to age 15, and combined mother and infants, central scenario

| Outcome and time horizon | Incremental | ICER (£) |
|---|---|---|
| **Expected cost (£)** | | |
| Mother | -55.84 | |
| Offspring | -135.65 | |
| Combined | --267.08 | |
| **LYGs** | | |
| Mother | 0.017 | -3220.55 |
| Offspring | 0.086 | -1581.98 |
| Combined | 0.103 | -2590.98 |
| **QALYs** | | |
| Mother | 0.054 | -1036.08 |
| Offspring | 0.098 | -1381.02 |
| Combined | 0.152 | -1,755.80 |
| **Benefit-cost ratio** | | |
| Mother | | 1.74 |
| Offspring | | 2.74 |
| Combined | | 4.31 |

### Supplementary Figure 1: Cost-effectiveness scatterplots for three main scenarios

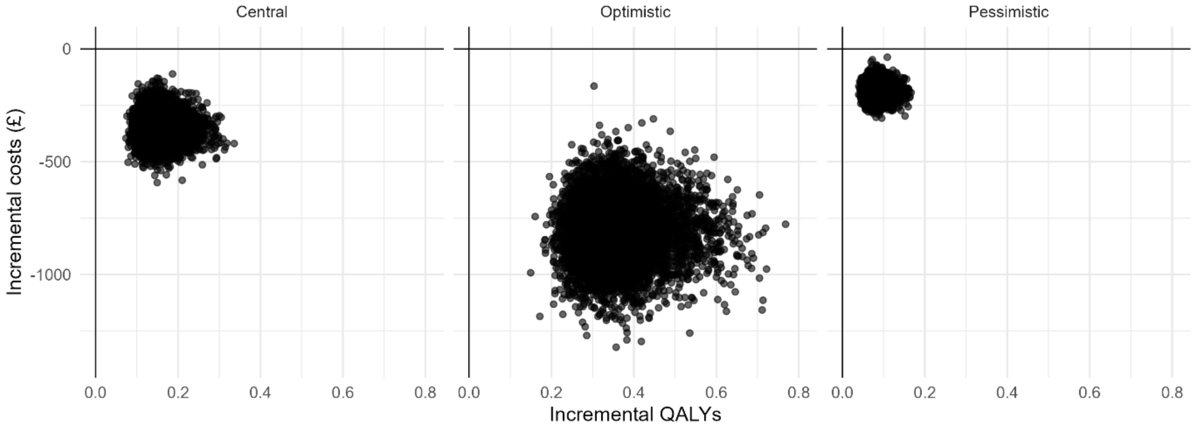
